# Evaluating the kidney disease progression using a comprehensive patient profiling algorithm: A hybrid clustering approach

**DOI:** 10.1101/2024.09.08.24313275

**Authors:** Mohammad A. Al-Mamun, Ki Jin Jeun, Todd Brothers, Ernest Asare, Khaled Shawwa, Imtiaz Ahmed

## Abstract

**Background:** Among 35.5 million U.S. adults with chronic kidney disease (CKD), more than 557,000 are on dialysis with incurred cost ranges from $97,373 to $102,206 per patient per year. Acute kidney injury (AKI) can lead to an approximate ninefold increased risk for developing CKD. Significant knowledge gaps exist in understanding AKI to CKD progression. We aimed to develop and test a hybrid clustering algorithm to investigate the clinical phenotypes driving AKI to CKD progression.

**Methods:** This retrospective observational study utilized data from 90,602 patient electronic health records (EHR) from 2010 to 2022. We classified AKI into three groups: Hospital Acquired AKI (HA-AKI), Community Acquired AKI (CA-AKI), and No-AKI. We developed a custom phenotypic disease and procedure network and a complementary variable clustering to examine risk factors among three groups. The algorithm identified top three matched clusters.

**Results:** Among 58,606 CKD patients, AKI group had a higher prevalence of heart failure (21.1%) and Type 2 Diabetes (45.3%). The No-AKI group had a higher comorbidity burden compared to AKI group, with average comorbidities of 2.84 vs. 2.04; p < 0.05; 74.6% vs. 53.6%. Multiple risk factors were identified in both AKI cohorts including long-term opiate analgesic use, atelectasis, history of ischemic heart disease, and lactic acidosis. The comorbidity network in HA-AKI patients was more complex compared to the No-AKI group with higher number of nodes (64 vs. 55) and edges (645 vs. 520). The HA-AKI cohort had several conditions with higher degree and betweenness centrality including high cholesterol (34, 91.10), chronic pain (33, 103.38), tricuspid insufficiency (38, 113.37), osteoarthritis (34, 56.14), and removal of GI tract components (37, 68.66) compared to the CA-AKI cohort.

**Conclusion:** Our proposed custom patient profiling algorithm identifies AKI phenotypes based on comorbidities and medical procedures, offering a promising approach to identify early risk factors for CKD using large EHR data.

## 1. Introduction

It has been estimated that over 35.5 million U.S. adults have chronic kidney disease (CKD), yet 9 out of 10 are unaware of their disease(1,2). Acute kidney injury (AKI) is independently associated with acute morbidity, mortality, and long-term kidney disease(3,4). Hospital-acquired acute kidney injury (HA-AKI) is a heterogeneous syndrome and a common complication in acute care settings and within the community. Among critically ill adults, up to 60% may experience this potentially preventable organ insult(5). A recent study found that patients who underwent dialysis for HA-AKI should receive follow up care due to its association with reduced mortality and lower hospitalization rates(6). However, less than 20% of HA-AKI survivors and only 21-50% of those who underwent acute dialysis within one year of hospitalization received nephrology follow-up(7,8). In contrast, AKI acquired in the community, known as community-acquired AKI (CA-AKI) differs in risk factors, epidemiology, presentation, and impact compared to HA-AKI. A recent study of 734,340 hospital admissions reported that patients with HA-AKI had higher rates of in-hospital mortality (51.58% vs. 26.07%), longer average hospital stays (35.84 ± 34.62 days vs. 21.25 ± 22.35 days), and a greater need for dialysis during hospitalization (2.06% vs. 1.45%) compared to those with CA-AKI (9).

The association between the initial occurrence of AKI and long-term risk of CKD is multifactorial and complex(10). Assessing the risk of AKI and its acceleration toward CKD or end-stage renal disease (ESRD) is a key area of research, with significant work focusing on its severity, recurrence, etiology, and clinical biomarkers(3,11). Studies have shown that patients who experience AKI have an approximately ninefold increased adjusted risk of developing CKD (pooled adjusted hazard ratio [HR] 8.8, 95% CI 3.1–25.5) and a threefold increased adjusted risk of progressing to ESRD (pooled adjusted HR 3.1, 95% CI 1.9–5.0)(12). Additionally, a 2016 study estimated that the prevalence of recurrent AKI after a first episode could be as high as 25% (13). AKI also contributes to a higher incidence of cardiovascular disease, AKI recurrence, and progression to CKD(3,9,14–16). However, identifying those at the highest risk, quantifying the extent of renal damage, and predicting the rate of disease progression remains challenging, even with well-known associated comorbidities such as hypertension, diabetes mellitus, and pre-existing CKD. No study has fully explored the relationship between comorbidities, medical procedures following AKI, and long-term kidney injury. Therefore, improving clinical phenotyping after an AKI event and understanding the trajectory toward CKD remains an area requiring further exploration and validation.

To better understand the multifactorial and complex progression, it is crucial to group clinical phenotypes and find the trajectory towards CKD which warrant a robust and comprehensive clustering methods. Traditional patient-centric clustering techniques, such as consensus clustering(17,18), K-means (19,20), latent class analysis (LCA) (21), and hierarchical clustering (22), often overlook the underlying data structure and complex interactions among variables, as they primarily focus on grouping individuals based on observed similarities. To effectively identify different phenotypes, it is essential to group variables (23–25) based on inherent correlations and dependencies, which allows fora more granular understanding of data structure and the extraction of latent pattern. Furthermore, representing comorbidity relationships as a network can help characterize pathways of disease progression to CKD by quantifying and analyzing pairwise interactions among variables (26). Current approaches are also limited in their ability to compare clinical phenotypes across multiple groups and are generally constrained by small datasets(23–26). Moreover, recent machine learning prediction models did not also consider a patient’s comprehensive phenotypic trajectory to predict AKI and its long-term outcomes (27–29).

To address this, we propose a novel phenotyping framework that employs two complementary methods: variable clustering and network-based clustering. We hypothesize that identifying high-risk clinical phenotypes through a comprehensive patient profiling algorithm will enhance our understand of factors associated with the progression of AKI to CKD. Variable clustering uncovers correlated clinical and demographic variables, revealing underlying patterns that influence disease progression. In contrast, network-based clustering constructs networks of variables, offering both visual and analytical insights to identify and group critical comorbidities and procedures that drive the progression from AKI to CKD. These two approaches complement each other by cross-referencing identified clusters, ensuring robust and accurate phenotyping. By integrating these methods and analyzing a large EHR dataset, our study aims to provide a comprehensive understanding of the factors driving AKI to CKD progression, thereby facilitating improved clinical decision-making and patient care. Additionally, this research contrasts phenotypes across distinct AKI subpopulations (HA-AKI, CA-AKI) to identify disease-specific trajectories.

## 2. Methods

### 2.1 Study Design and Setting

This was a single center, STROBE-compliant retrospective observational study involving 90,602 patients with records from February 2010 to 2022. Given the retrospective nature of the data, informed consent was not required, as all patient information was de-identified prior to use. The study received an exemption from the Human Research Review Committee at the West Virginia University Institutional Review Board (IRB: # 2212689753)

### 2.4 Data sources

Data were obtained from the electronic health records (EHR) of a single Health Care Organization (HCO), namely, West Virginia University (WVU), using TriNetX on 13/06/2022. TriNetX is a global health research network that connects pharmaceutical companies, study sites, investigators and patients by sharing real-world data to facilitate clinical and observational research. The dataset typically includes information on diagnoses, procedures, encounters, medications, laboratory results, vital signs, genomic data, tumor properties, oncology treatments, tumor, chemotherapy lines, cohort details, and demographics.

### 2.2 Participants

Patients aged 18 years and older diagnosed with CKD using ICD-9-CM code 585 and ICD-10-CM code N18 were included in the study. These patients were followed retrospectively from the inception of the database until September 2022.

### 2.3 Definitions, Inclusion, and Exclusion criteria

Prior AKI events were identified using ICD-9-CM code 485 and ICD-10-CM code N17. CKD patients were followed for three years to determine the incidence of AKI. Patients with AKI events within three years prior to their CKD diagnosis or with any dialysis events before their CKD diagnosis were excluded from the study. HA-AKI patients were defined as those who had AKI within 90-days of any inpatient hospitalization (identified by Current Procedural Terminology (CPT) codes: 99218-99239, 99251-99255, 99291, 99304-99307, 94002, G03378,1013699, and 1013659). CA-AKI patients were defined as those who had any AKI events, excluding HA-AKI, within three years prior their CKD diagnosis. After applying the inclusion and exclusion criteria, we created three cohorts: HA-AKI, CA-AKI, and No-AKI. A study design flow chart is shown in Fig 1.

**Fig 1:**
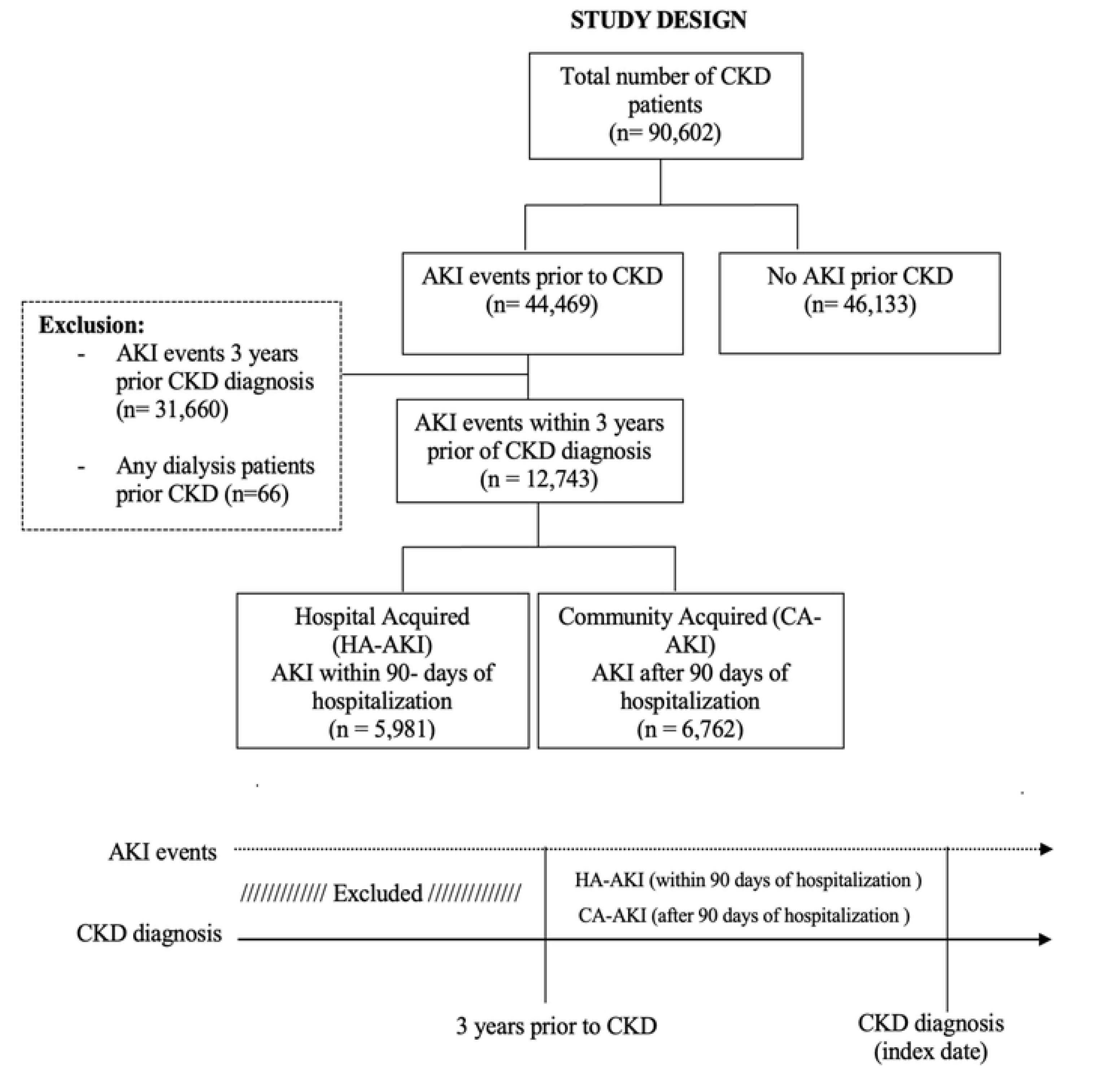
Study design schema for the three study cohorts and their timelines. Abbreviations: CKD-Chronic Kidney Disease, HA-AKI-Hospital acquired acute kidney injury, and CA-AKI-Community acquired acute kidney injury.

### 2.7 Identification of Comorbidities and Procedures

We examined and ranked all diagnoses and procedures codes for three cohorts. To obtain more reliable estimates, we excluded rarely diagnosed diseases and conducted procedures with a prevalence of less than 1% across all cohorts. Before ranking, we converted ICD-CM 9 codes to ICD-CM 10 codes to ensure consistency across cohorts. For the HA-AKI cohort, there were 64 diagnosis codes and 62 procedures codes. For the CA-AKI cohort, there were 62 diagnosis codes and 65 procedure codes. For the No-AKI cohort, there were 55 diagnoses codes and 67 procedure codes. Supplementary Tables 1 and 2 provide the ICD-10-CM codes and combined CPT codes, along with their prevalence for each cohort, respectively.

### 2.6 Statistical Analysis

Descriptive statistics were employed to characterize the study population with continuous variables reported as means and standard deviations and categorical variables described using frequencies and proportions. Baseline characteristics of HA-AKI patient were compared with No-AKI patients, and CA-AKI were compared with No-AKI patients using Pearson chi-square tests for categorical variables and independent-samples t-tests for continuous variables.

### 2.8 Comorbidity network modeling

To understand the risk factor profiles associated with HA-AKI, CA-AKI, and non-AKI patients in relation to CKD, we constructed phenotypic disease and procedure network models. We binarized all diagnoses and procedures for each cohort based on their presence or absence prior to CKD diagnosis. For procedures, we limited the analysis to those occurring within three years before CKD diagnosis. Phenotypic networks were then created for each cohort, with nodes representing disease diagnoses (comorbidities) and edges indicating co-occurrence relationships between pairs of comorbidities(26,30,31). Network analysis offers a graphical representation of the complex patterns among risk factors. To quantify the strength of comorbidities between AKI to CKD and No-AKI to CKD groups, we introduced the observed-to-expected ratio (OER)(32). The OER_ij_, measures the strength of the comorbidity between disease pair *i* and *j*, calculated as the ratio of the observed prevalence of the disease pair (O) to the expected prevalence (E), which is determined by the product of the prevalence of diseases *i* and *j*:

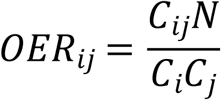

In this context, C_ij_ represents the co-occurrence count of both diseases (*i* and *j*), *N* is the total number of patients in the population, and C_i_ and C_j_ are the prevalence of diseases (*i* and *j*). The observed-to-expected ratio (OER) can be interpreted as a measure of relative risk. An OER greater than1 indicates that the two diseases co-occur more frequently in the same patients than would be expected by chance, whereas an OER less than 1 suggests that the diseases are mutually exclusive. We included only those pairwise comorbidities in the network with a OERs greater than 1, as we aimed to identify risk factors specific to each cohort. To highlight prominent risk factors in the network graphs, we set the 90^th^ percentile as the threshold for each cohort. For diagnoses, OER thresholds were 1.68, 1.84, and 2.25 for the HA-AKI, CA-AKI, and No-AKI cohorts, respectively. For procedures, the OER thresholds were 2.08, 2.14, 2.75 for the HA-AKI, CA-AKI, and No-AKI cohorts, respectively.

To explore the complexity among the diagnoses and procedures, we incorporated five network metrics: diameter, degree centrality, betweenness centrality, average nearest neighbor path length, and closeness centrality (33). Degree centrality represents the number of direct connections a node has with other diseases or procedures. An edge in the network signifies the interaction or relationship between nodes. The average degree is the mean number of edges connected to each node. The diameter of the network indicates the maximum number of edges that must be traversed to travel between the most distant nodes. Betweenness centrality measures the extent to which a node lies on the shortest paths between other pairs of nodes in the network. Closeness centrality is the reciprocal of the sum of the shortest path lengths between a node and all other nodes in a network. A higher closeness centrality value indicates that a disease is more likely to be diagnosed with other diseases in fewer steps. We used an unweighted edge to represent the presence or absence of relationships in a network, which means the network lacks temporal directionality for diseases and procedures. To identify cluster of closely related comorbidities, we employed the fast-unfolding cluster algorithm for community detection, which optimizes for the highest modularity score across different clustering layers (34,35). This analysis allowed us to identify the top three clusters of diagnoses and procedures for each cohort. Network analysis was performed using the *Gephi* network software package (36).

### 2.9 Hierarchical Diagnosis and Procedure Clustering

To validate the community clustering of variables from the phenotypic network analysis, we employed a variable clustering method known as “*ClustOfVar*“ (37). This advanced algorithm is well-suited for datasets containing both quantitative and qualitative variables (37). ClustOfVar provides a comprehensive understanding of data structure by grouping variables into clusters based on their similarities. It identifies groups of strongly related variables and constructs synthetic variables that summarize these clusters, thereby reducing data complexity while preserving key information. The primary metric used in *ClustOfVar* is the *homogeneity criterion*, which assesses how closely related the variables within a cluster are to a synthetic variable representing that cluster. For quantitative variables, homogeneity is measured by the squared Pearson correlation, while for qualitative variables, it is measured by the correlation ratio. Mathematically, the homogeneity *H*(*C*_*k*_) for a cluster *C_k_* is defined as:

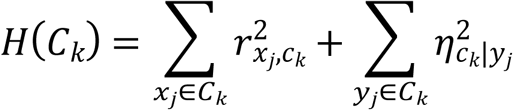

Where r*_xj,ck_*^2^ is the squared Pearson correlation between quantitative variable *x*_*j*_ and the synthetic variable *c*_*k*_ and *η^2^_ck|yj_* is the correlation ratio between qualitative variable *y*_*j*_ and the synthetic variable *c*_*k*_. The synthetic variable *c*_*k*_ is created using principal component analysis (PCA) for quantitative variables and multiple correspondence analysis (MCA) for qualitative variables. This synthetic variable represents the first principal component derived from applying PCA or MCA to all variables within the cluster. We utilized a hierarchical clustering approach to perform the clustering process. This method begins by treating each variable as an individual cluster and iteratively merges clusters to minimize the loss of homogeneity. The dissimilarity between any two clusters *C*_1_and *C*_2_is defined as:

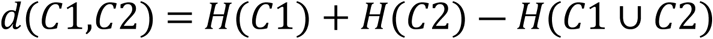

The algorithm continues to merge clusters until all variables are consolidated into a single cluster, resulting in a dendrogram. This dendrogram can be cut at different levels (*k*) to achieve the desired number of clusters. The hierarchical approach provides a clear visual representation of the clustering process, helping to determine the optimal number of clusters. In summary, the *ClustOfVar* algorithm offers a robust framework for variable clustering by maximizing homogeneity within clusters. It employes advanced statistical methods to handle mixed data types, ensuring a comprehensive understanding of the underlying data structures and facilitating the identification of meaningful patterns. Supplementary file 1 outlines the steps of the *ClustOfVar* algorithm in Step II. For further details, please refer to Chavent et al.(25)

Each method identified the top three clusters for diagnoses and procedures within each cohort. We then matched these clusters and calculated the similarity percentage among them. The final selection of diagnosis and procedure variables was based upon these matches. We anticipated that the hierarchical clustering algorithm would yield similarly complex networks of variables as those identified by the phenotypic disease network. Details of the algorithms used for comprehensive patient profiling and cluster matching between the two methods are provided in Supplementary File 1.

## 3. Results

### 3.1 Demographic and Clinical Characteristics

A total of 58,606 patients were included in the analysis. The sample predominantly comprised White (84.02%), non-Hispanic or Latino (82.87%), females (52.03%) with a mean age of 61 years. Table 1 outlines the demographic characteristics of individuals with CKD who experienced AKI compared to those who did not. The AKI group had a higher proportion of White (88.7%) or non-Hispanic/Latino individuals (90.0%), a slightly higher percentage of females (51.4%), and a lower mean age 58.1 years) compared to the No-AKI group (61.6 years). The AKI group had a significantly lower mean age (65.8 ± 13.4 years) compared to the No-AKI group (68.6 ± 13.4 years). Additionally, the No-AKI group had a higher comorbidity burden, with a larger proportion having at least two comorbidities (mean number of comorbidities = 2.84 vs. 2.04; p < 0.05; 74.6% vs. 53.6%, respectively). Among the No-AKI group, 76.3% had hypertension, 33.7% had nicotine dependence, and 38.8% had Type 2 Diabetes Mellitus. In contrast, the AKI group had a higher prevalence of heart failure (21.1%) and Type 2 Diabetes Mellitus (45.3%).

**Table 1:**
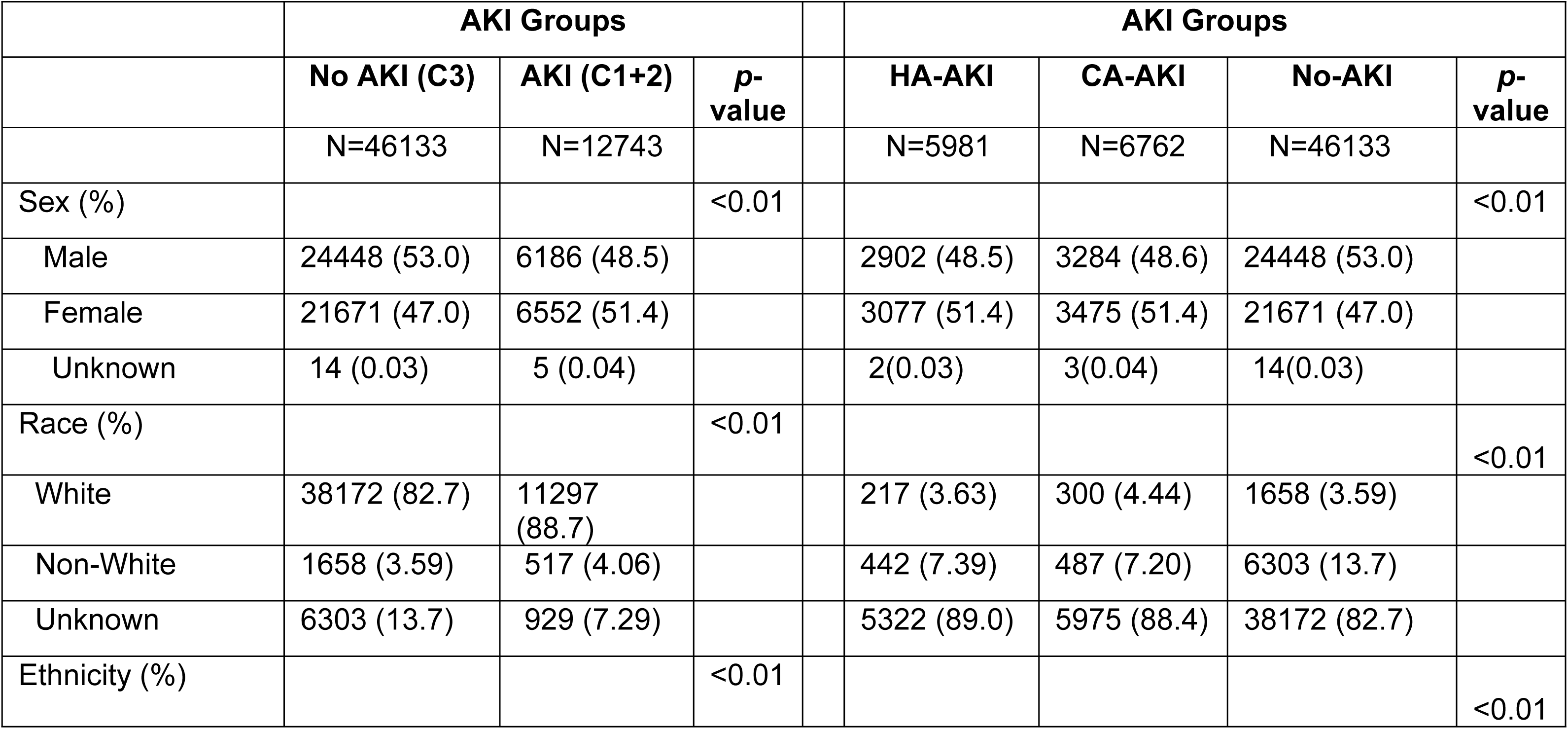
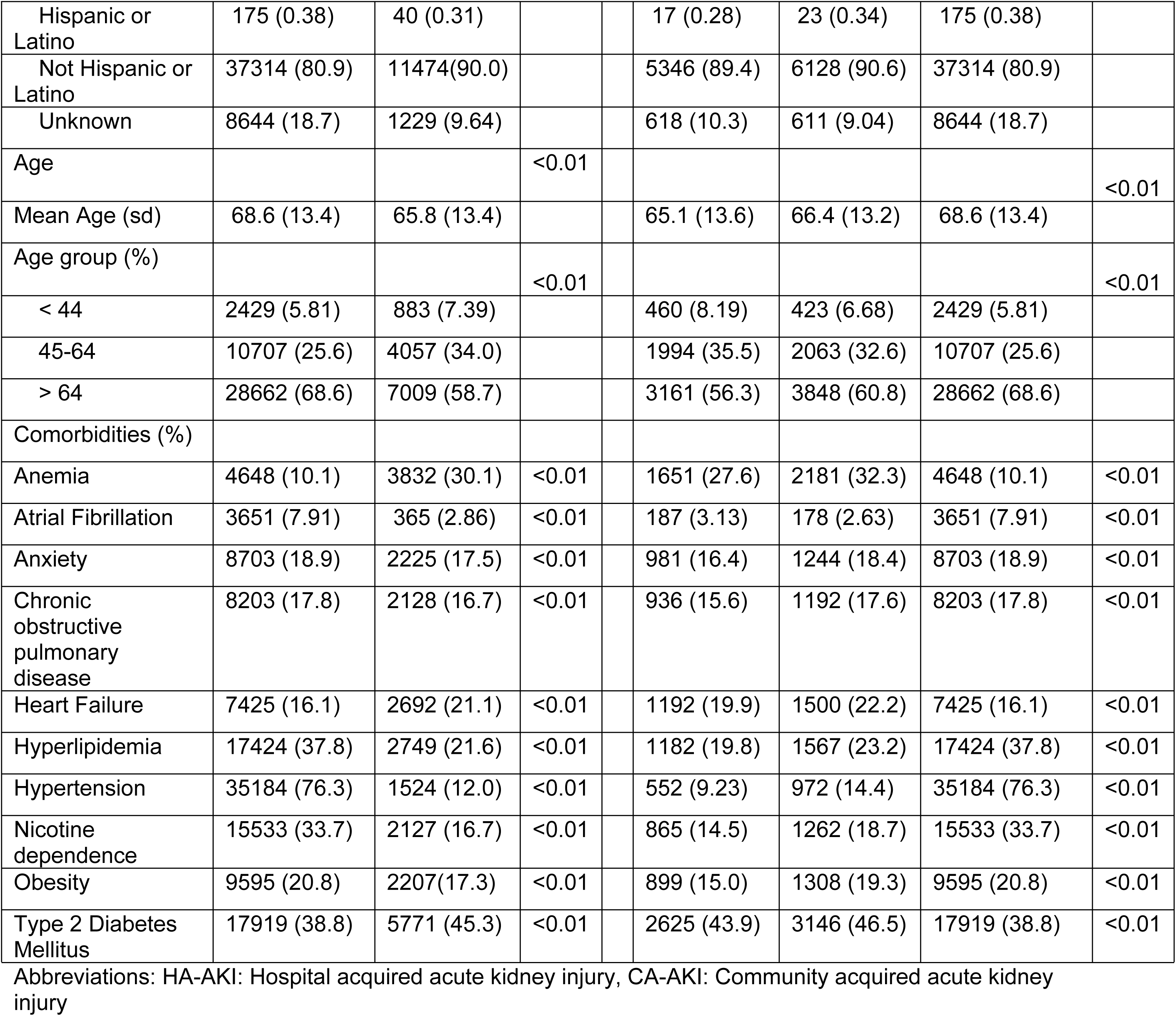
Demographics characteristics of AKI vs No-AKI and among the groups.

### 3.2. Network analytics of comorbidity and procedures

**Fig 2a** represents the visual network connections of study participants’ diagnoses using ‘betweenness centrality’ as the ranking factor to identify key nodes acting as critical ‘bridges’ between components within the network. Nodes with high ‘betweenness’ are strategic targets for control. OER values greater than 1 were selected to highlight the strength of relationships between variables, with higher OERs indicating stronger associations. Multiple nodes (ICD-9/10 codes) were identified in cohorts 1 and 2, but not cohort 3. Notably, the key nodes in cohorts 1 and 2 included: Z79.891 (long-term opiate analgesic use), J98.11 (atelectasis), Z82.49 (history of ischemic heart disease), E87.2(lactic acidosis) and E11.65(Type 2 diabetes mellitus with hyperglycemia). In contrast, the most significant nodes in the control group were I25.2, (previous myocardial infarction), Z90.49 (absence of other specified parts of digestive tract), and Z82.49 (family history of ischemic heart disease).

**Supplementary Fig 1.**
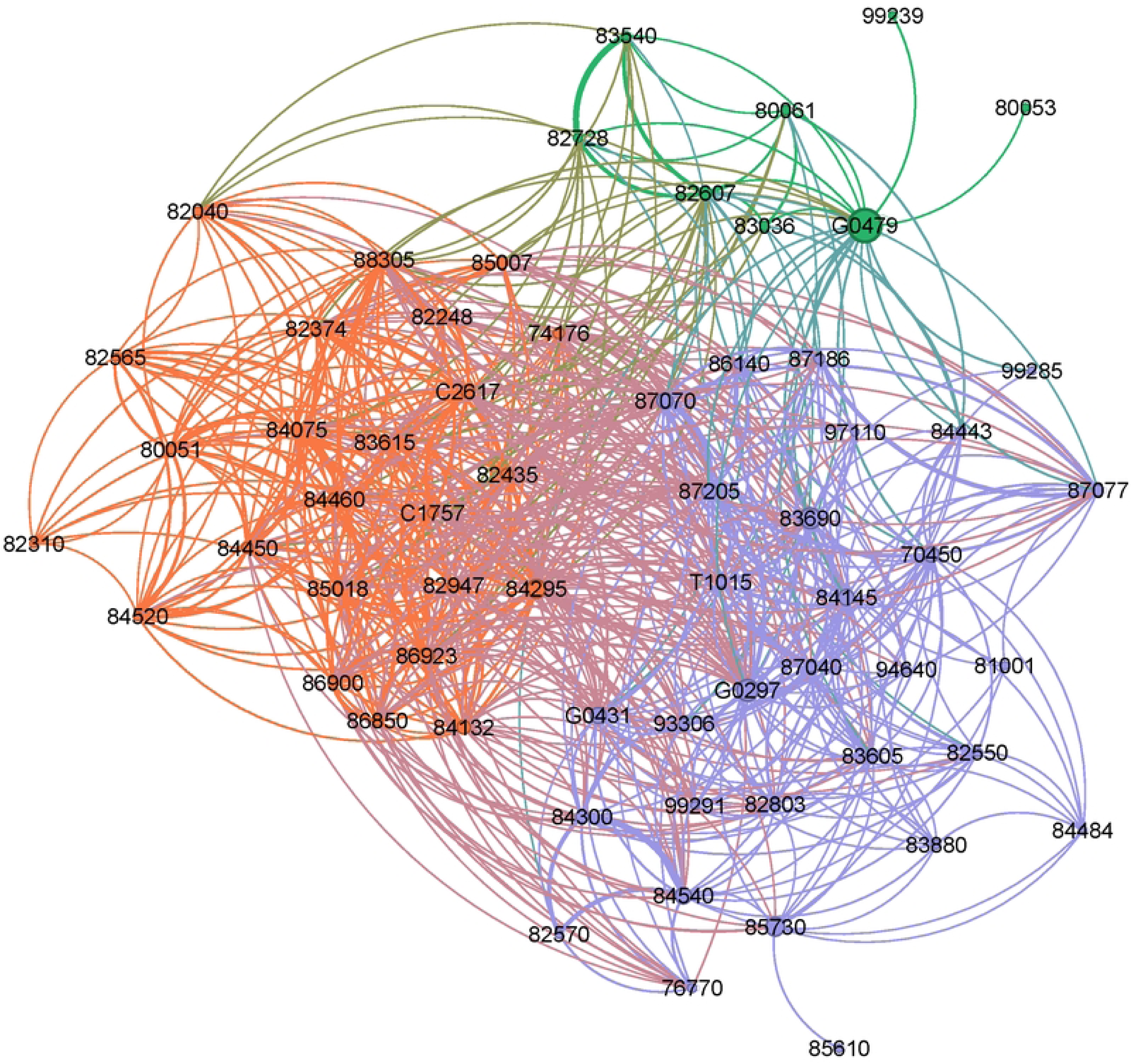
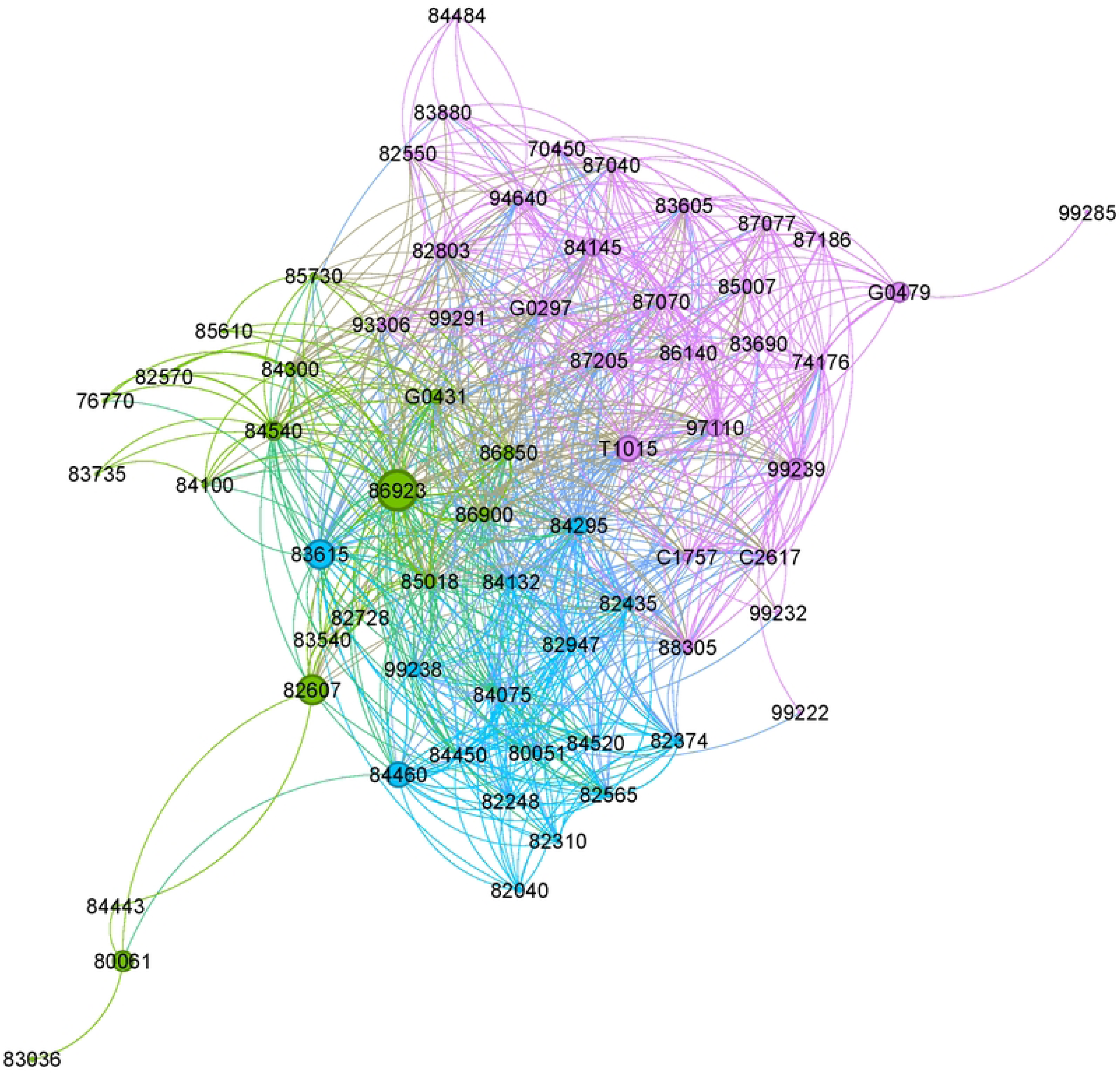
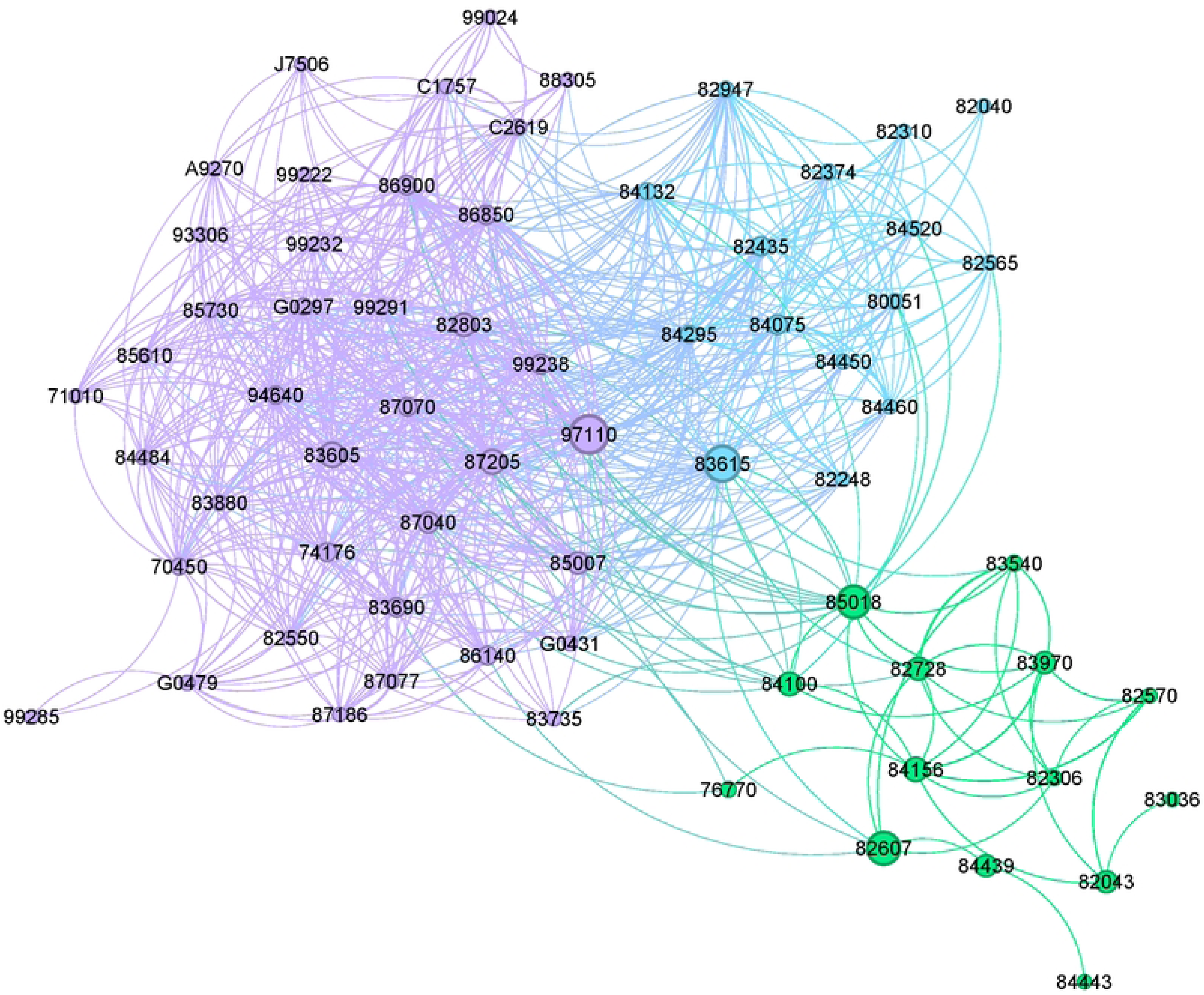
depicts the network of procedures encountered across the three cohorts (HA-AKI, CA-AKI, and no-AKI). Several nodes were common to both the HA-AKI and CA-AKI cohorts, including G0479 (drug test(s), 84145 (procalcitonin level evaluation), 84295 (sodium concentration measurement), 84540 (urea nitrogen measurement), and 86923 (transfusion cross-matching donor blood compatibility). Unique to both the control and AKI patient networks were 82607 (vitamin B12measurement) and 83615 (lactate dehydrogenase measurement). Nodes specific to cohort 3 (No-AKI) included 83605 (serum lactate measurement) and 85018 (hemoglobin measurement in whole blood samples).

**Fig 2:**
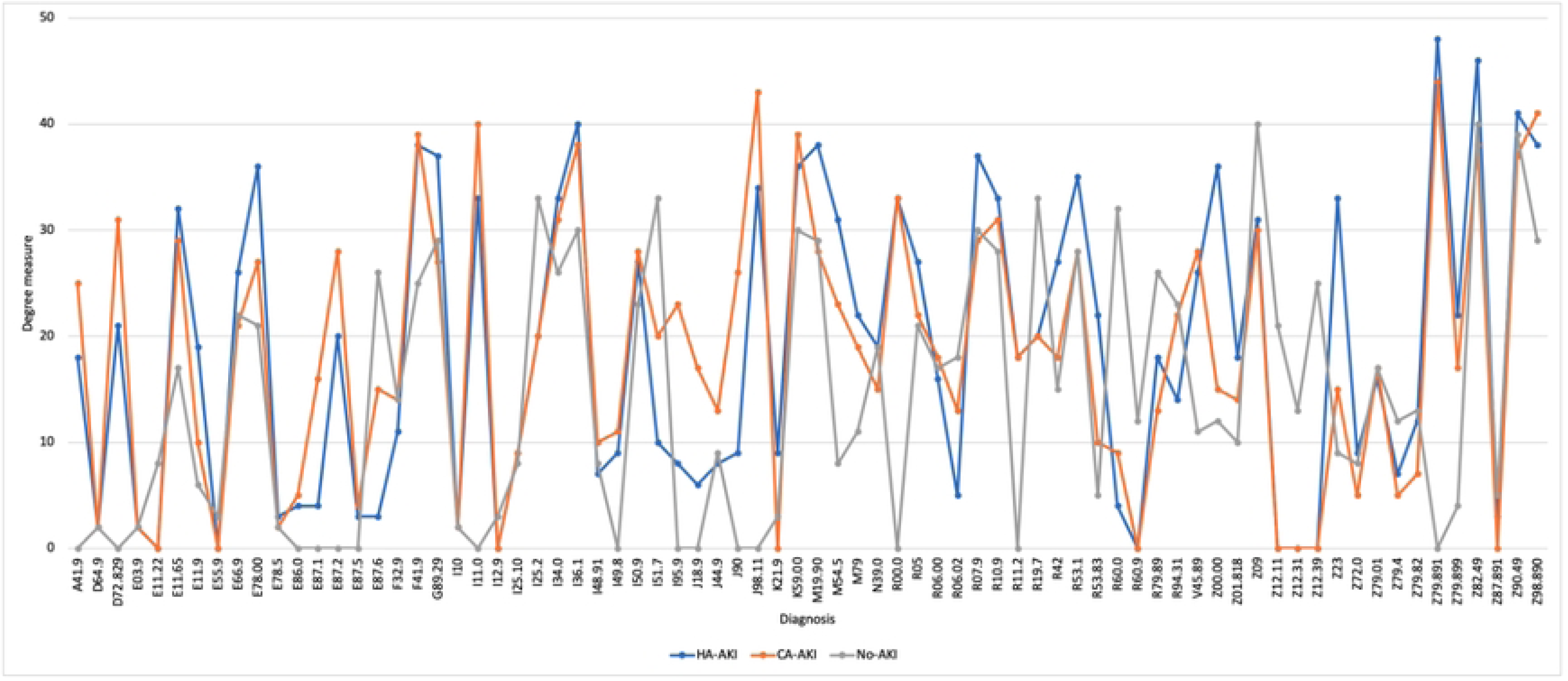
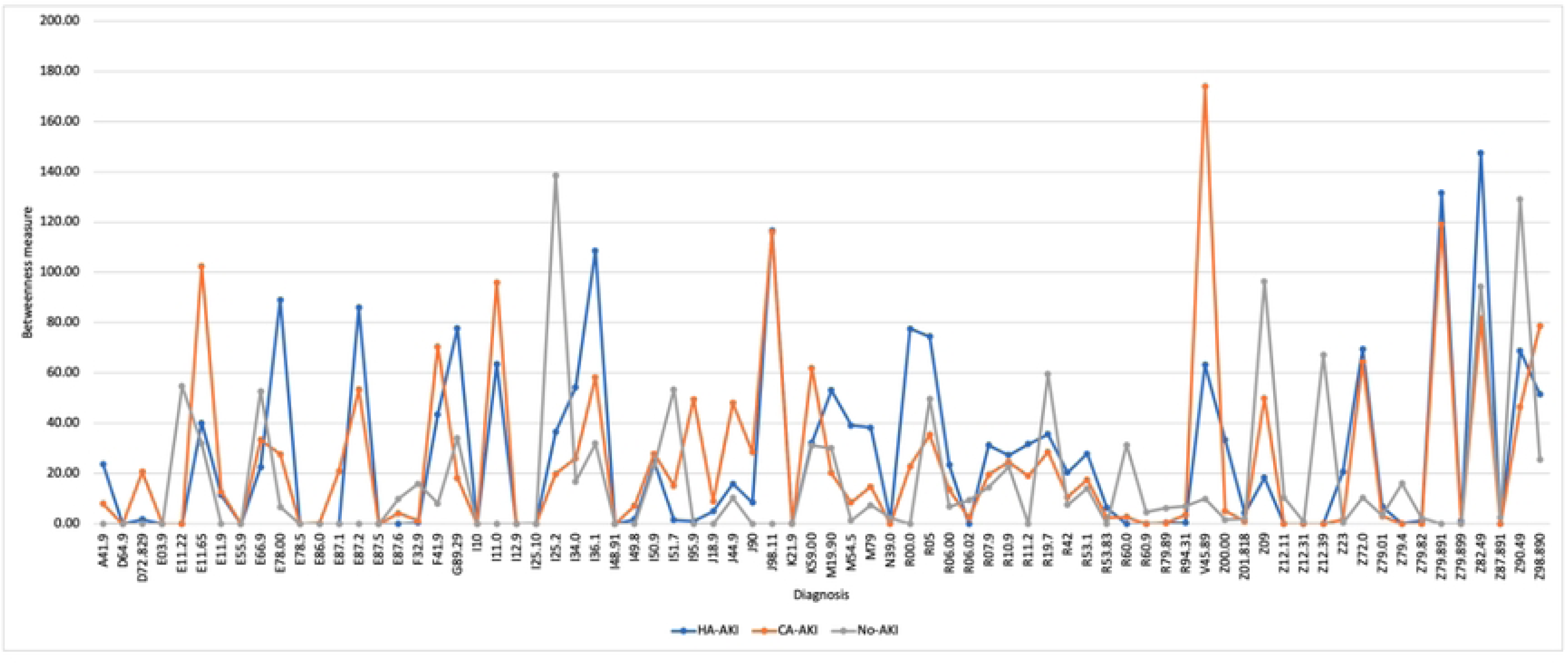
The strongest comorbidity associations across the three cohorts: a) HA-AKI, b) CA-AKI, and c) No-AKI. Nodes represent comorbidities and are color-coded by disease clusters. Node size reflects prevalence, while edge thickness indicates the observed-to-expected ratio (OER). Only OER values within the 90^th^ percentile are displayed.

**Supplementary Fig 2.** The strongest associations of medical procedures across the three cohorts: a) HA-AKI, b) CA-AKI, and c) No-AKI. Nodes represent procedures and are color-coded by procedure clusters. Node size indicates prevalence, while edge thickness represents the observed-to-expected ratio (OER). Only OER values within the 90^th^ percentile is shown.

**Table 2** provides an overview of the network parameters for diagnoses and procedures across the three cohorts. While the complexity of the procedure networks was similar among the cohorts, the comorbidity network in HA-AKI patients was more complex compared to the No-AKI group, as evidenced by the higher number of nodes (64 vs. 55) and edges (645 vs. 520). The diagnosis network in the HA-AKI cohort was also more tightly clustered compared to the No-AKI cohort, as indicated by a significantly higher average degree (21.68 vs. 16.91, p < 0.04). For procedures, both AKI groups showed significantly higher measures for average degree, path length, betweenness and closeness compared to the No-AKI group.

**Table 2.**
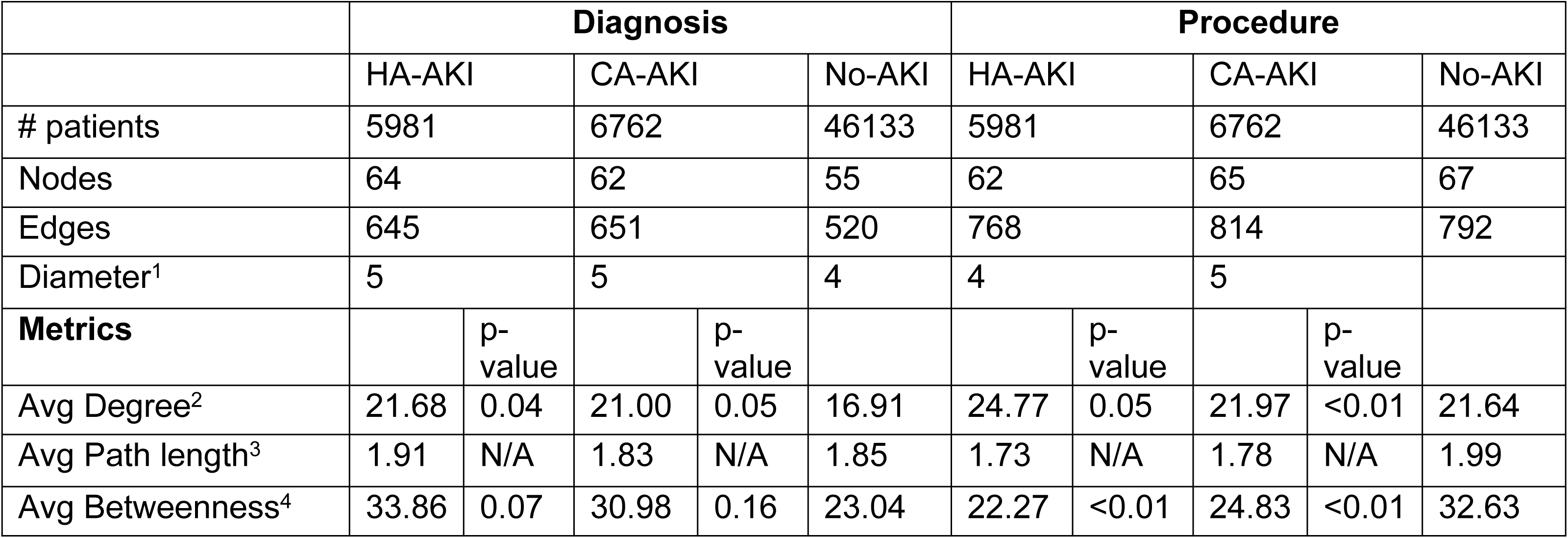
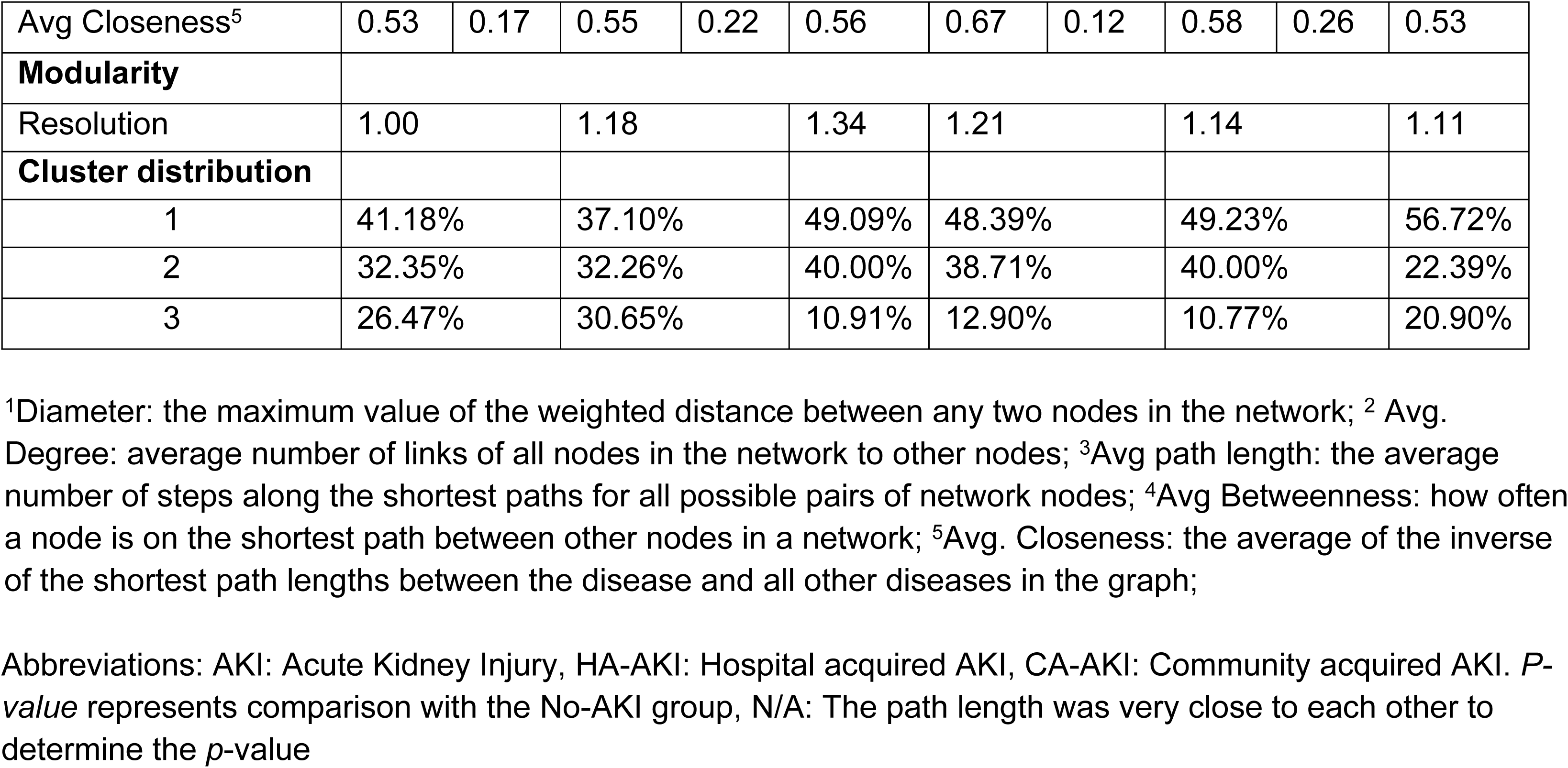
Network metrics in the comorbidity network of HA-AKI, CA-AKI, and No-AKI patients.

Fig 3 illustrates the average degree and betweenness measures of the comorbidity network for three cohorts. A higher degree value in one cohort suggests a greater number of connections between nodes, indicating a higher comorbidity burden. Higher betweenness centrality represents the overlap of comorbid conditions, as shown in Fig 3.

**Fig 3.**
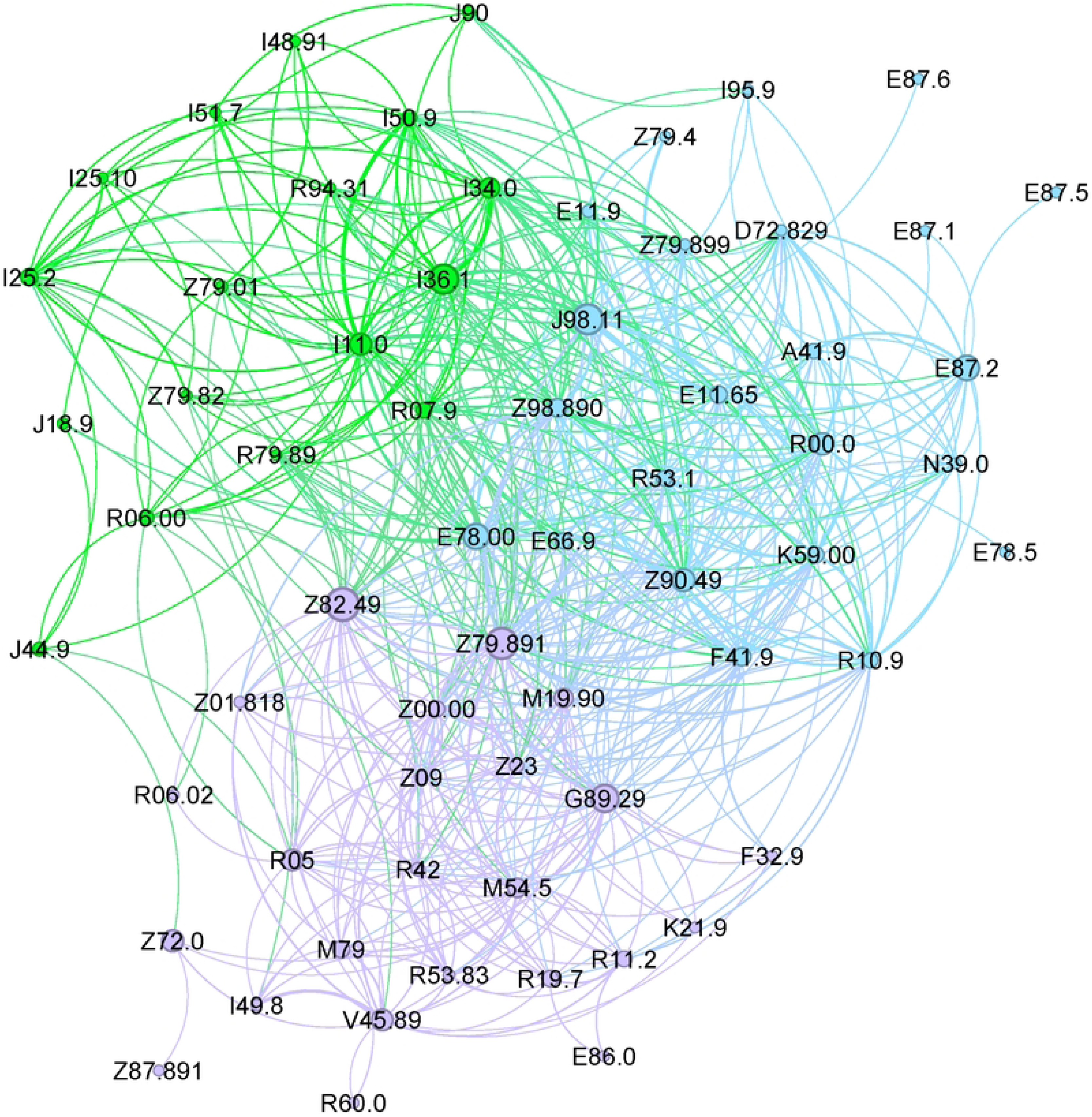
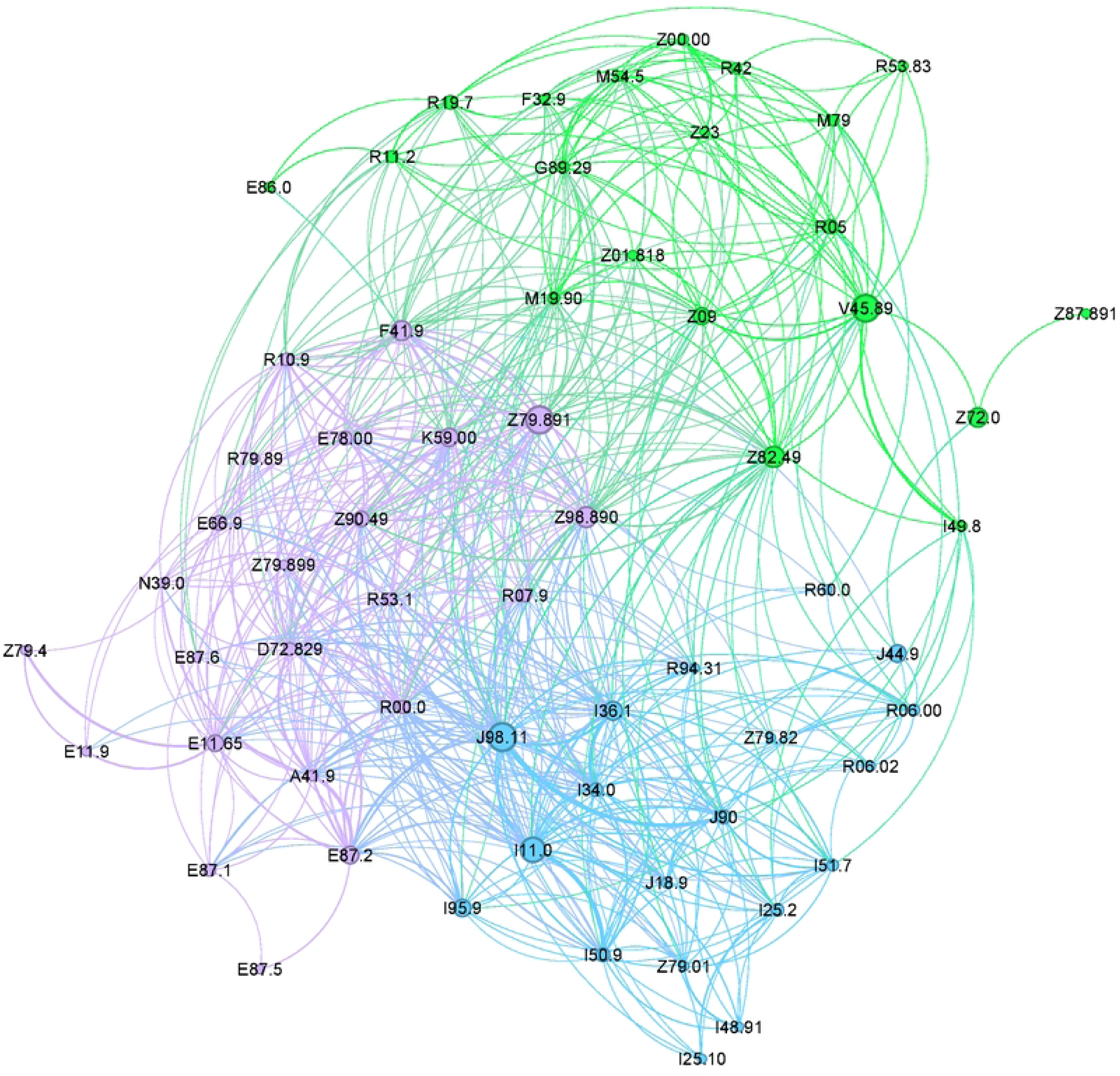
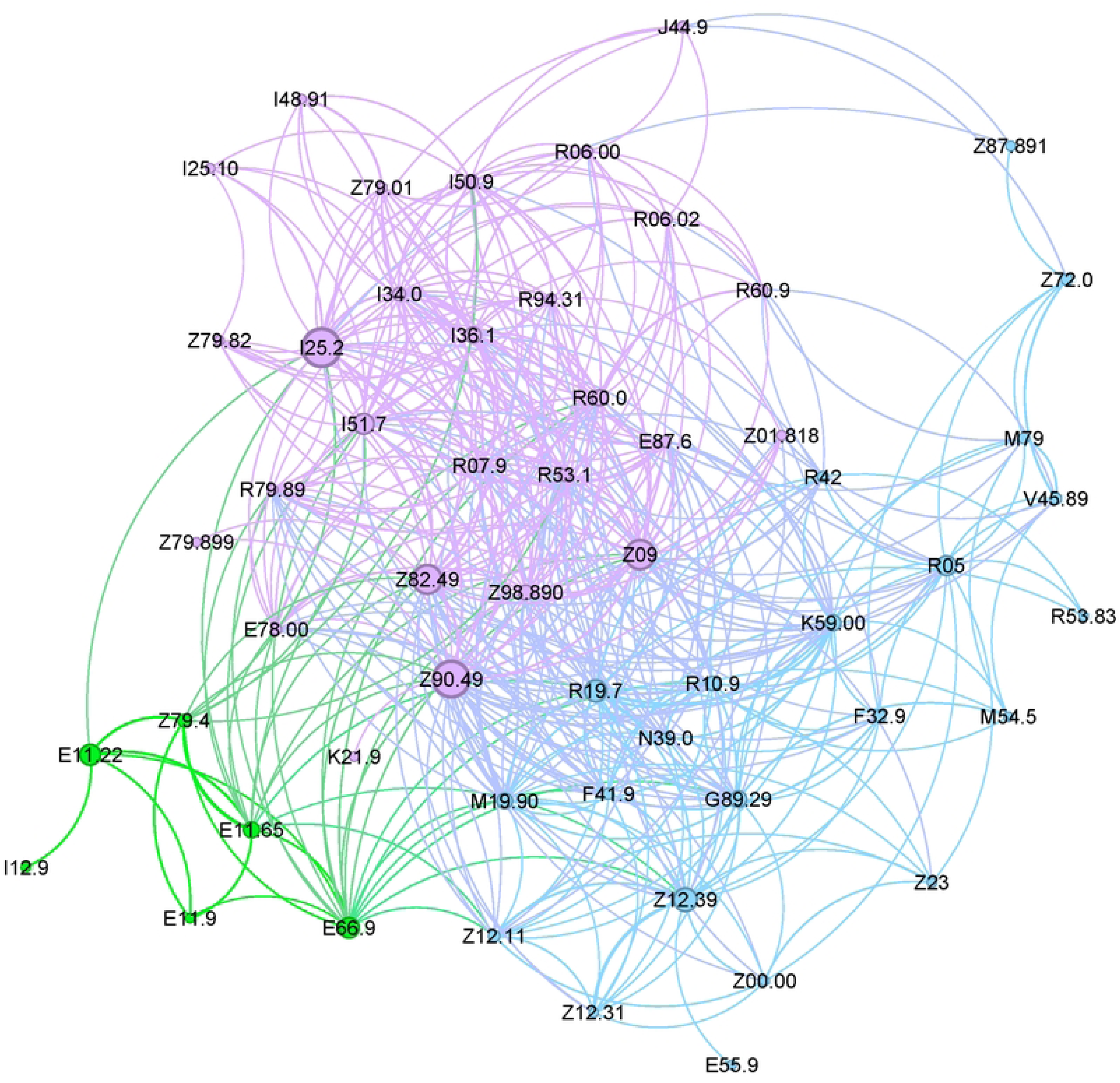
Centrality measures using a) degrees and b) betweenness among the three cohorts (HA-AKI, CA-AKI, No-AKI) for comorbidities.

### 3.3 Matched phenotypes

Our custom matching algorithm yielded the best alignment of clusters between both methods. Supplementary Table 3 shows the percentage of clusters matched by both algorithms. For HA-AKI and CA-AKI, we achieved a 96% match among commodity phenotypes, while for the No-AKI cohort, the match was 100%. Table 3 presents the similar phenotypes found across the three cohorts. Among these cohorts, we identified 10 cardiovascular-related conditions, including hypercholesterolemia, atherosclerotic heart disease without angina pectoris, myocardial infarction, non-rheumatic mitral insufficiency, nonrheumatic tricuspid insufficiency, atrial fibrillation, heart failure, cardiomegaly, chest pain, and abnormal ECG/EKG findings. However, the *degree* and *betweenness* centrality differ across the cohorts.

**Table 3.**
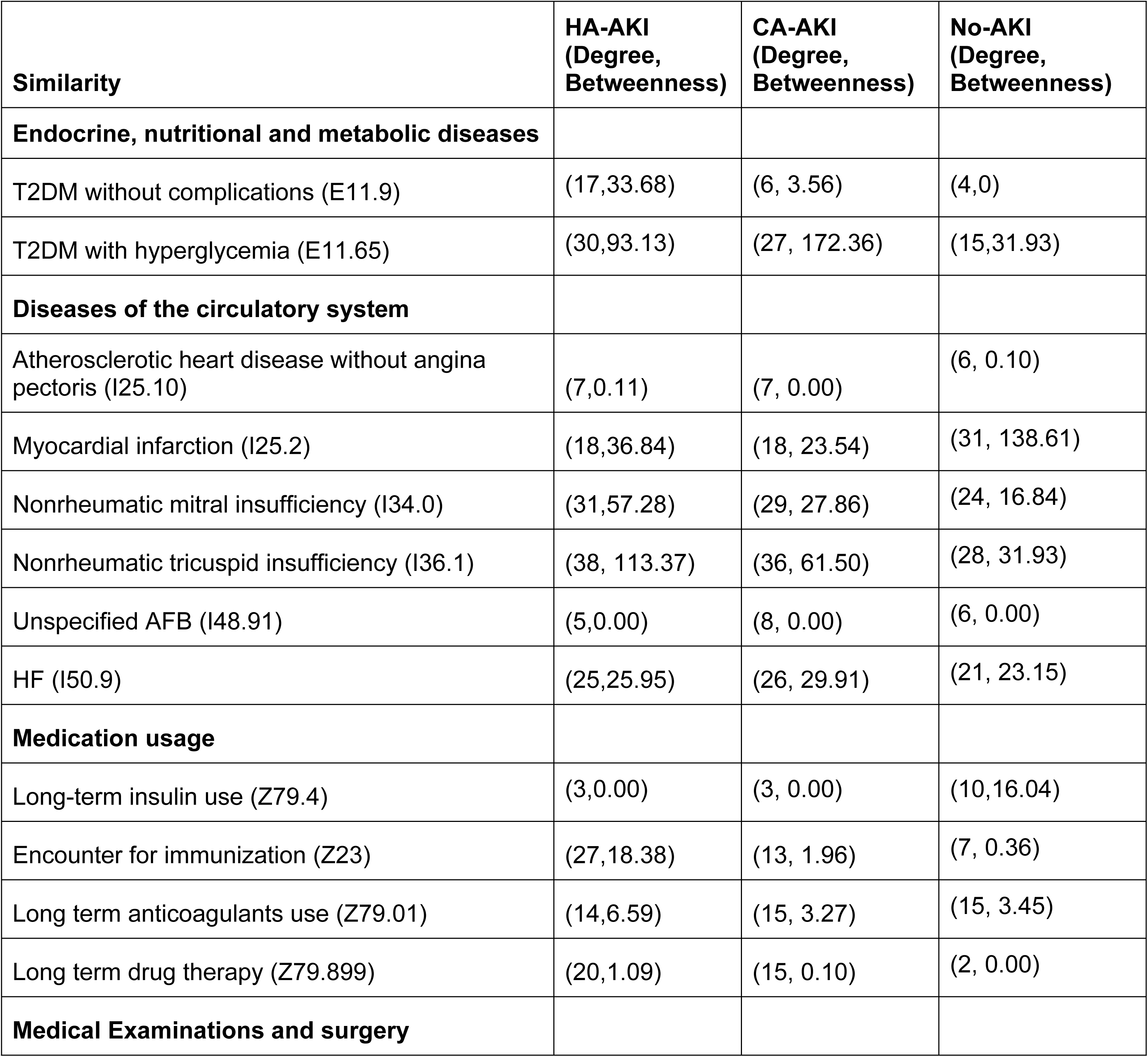
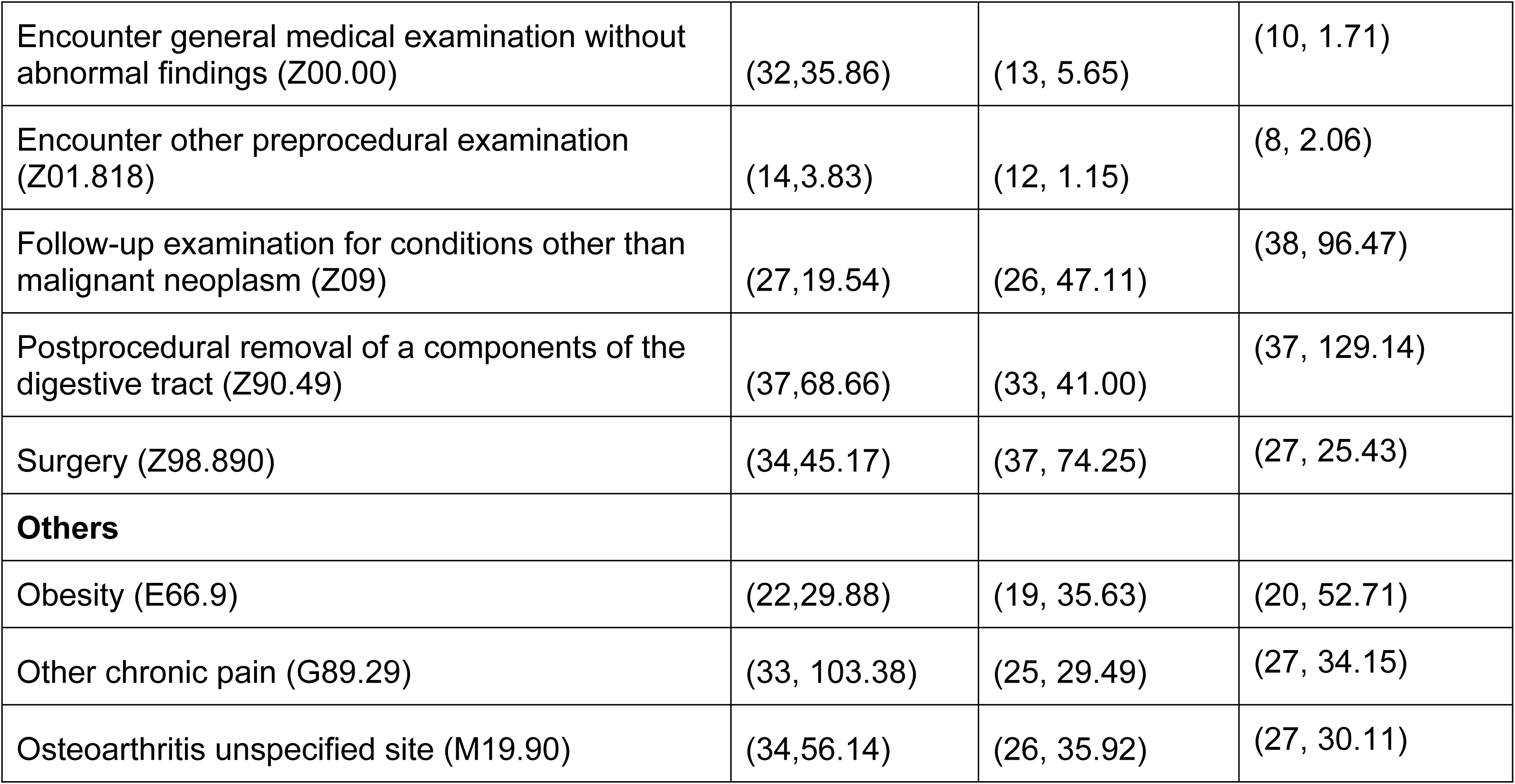
Comparison of diagnoses across the three cohorts. Similarities were determined by comparing the best-matched clusters identified by both algorithms. The table presents the degree and betweenness values for each comorbidity within the cohorts.

| Similarity | HA-AKI<br>(Degree,<br>Betweenness) | CA-AKI<br>(Degree,<br>Betweenness) | No-AKI<br>(Degree,<br>Betweenness) |
| --- | --- | --- | --- |
| **Endocrine, nutritional and metabolic diseases** |  |  |  |
| T2DM without complications (E11.9) | (17,33.68) | (6, 3.56) | (4,0) |
| T2DM with hyperglycemia (E11.65) | (30,93.13) | (27, 172.36) | (15,31.93) |
| **Diseases of the circulatory system** |  |  |  |
| Atherosclerotic heart disease without angina pectoris (I25.10) | (7,0.11) | (7, 0.00) | (6, 0.10) |
| Myocardial infarction (I25.2) | (18,36.84) | (18, 23.54) | (31, 138.61) |
| Nonrheumatic mitral insufficiency (I34.0) | (31,57.28) | (29, 27.86) | (24, 16.84) |
| Nonrheumatic tricuspid insufficiency (I36.1) | (38, 113.37) | (36, 61.50) | (28, 31.93) |
| Unspecified AFB (I48.91) | (5,0.00) | (8, 0.00) | (6, 0.00) |
| HF (I50.9) | (25,25.95) | (26, 29.91) | (21, 23.15) |
| **Medication usage** |  |  |  |
| Long-term insulin use (Z79.4) | (3,0.00) | (3, 0.00) | (10,16.04) |
| Encounter for immunization (Z23) | (27,18.38) | (13, 1.96) | (7, 0.36) |
| Long term anticoagulants use (Z79.01) | (14,6.59) | (15, 3.27) | (15, 3.45) |
| Long term drug therapy (Z79.899) | (20,1.09) | (15, 0.10) | (2, 0.00) |
| **Medical Examinations and surgery** |  |  |  |
| Encounter general medical examination without abnormal findings (Z00.00) | (32,35.86) | (13, 5.65) | (10, 1.71) |
| Encounter other preprocedural examination (Z01.818) | (14,3.83) | (12, 1.15) | (8, 2.06) |
| Follow-up examination for conditions other than malignant neoplasm (Z09) | (27,19.54) | (26, 47.11) | (38, 96.47) |
| Postprocedural removal of a components of the digestive tract (Z90.49) | (37,68.66) | (33, 41.00) | (37, 129.14) |
| Surgery (Z98.890) | (34,45.17) | (37, 74.25) | (27, 25.43) |
| **Others** |  |  |  |
| Obesity (E66.9) | (22,29.88) | (19, 35.63) | (20, 52.71) |
| Other chronic pain (G89.29) | (33, 103.38) | (25, 29.49) | (27, 34.15) |
| Osteoarthritis unspecified site (M19.90) | (34,56.14) | (26, 35.92) | (27, 30.11) |

Betweenness centrality measures the extent to which a node lies on the shortest paths between other nodes, indicating its influence within the network by controlling the flow of information. In the HA-AKI cohort, conditions like chronic pain and osteoarthritis had higher degree and betweenness values (33, 103.38; 34, 56.14) compared to CA-AKI (25, 29.49; 26, 35.92) and no-AKI (27, 34.15; 27, 30.11) cohorts, respectively.

When comparing HA-AKI to CA-AKI, several conditions exhibited higher betweenness centrality in the HA-AKI cohort, including high cholesterol (34, 91.10), chronic pain (33, 103.38), tricuspid insufficiency (38, 113.37), osteoarthritis (34, 56.14), general medical examination (32, 35.86), and removal of GI tract components (37, 68.66). Conversely, in the CA-AKI cohort, cardiomegaly (18, 14.83), abnormal ECG (20, 3.68), follow up exam (26, 47.11), and surgery (37, 74.25) showed higher betweenness values. When comparing either AKI cohort to the non-AKI group, the non-AKI cohort had higher betweenness centrality for conditions such as obesity (20, 52.71), myocardial infarction (31, 138.61), cardiomegaly (31, 53.45), abnormal chemistry findings (24, 6.30), follow up examinations (38, 96.47), and removal of components of the GI tract (37, 129.14).

Table 4. Dissimilarities Among the Three Cohorts. In comparing the degree and betweenness of diagnoses between the HA-AKI and CA-AKI cohorts, we observed that sepsis (16, 26.31), tachycardia (29, 68.27), and dizziness/giddiness (23, 20.92) exhibited higher values in the HA-AKI cohort. Conversely, in the CA-AKI cohort, diagnoses such as type 2 diabetes with hyperglycemia (27, 172.36), hypoosmolality and hyponatremia (14, 22.00), hypertensive heart disease with heart failure (38, 99.68), COPD (unspecified) (11, 52.62), elevated WBC count (29, 22.26), and pleural effusion (24, 27.98) showed higher degree and betweenness compared to the HA-AKI cohort. Interestingly, anxiety disorder (A41.9) was the second-highest node in the HA-AKI cohort, with values of 36 and 53.95, respectively. It is noteworthy that pleural effusion (J90) had minimal impact on the phenotypic network of the CA-AKI cohort. Additionally, certain non-traditional factors, such as unspecified constipation, unspecified abdominal pain, and other external hearing aids, exhibited high degree and betweenness in both AKI cohorts but were not detected by the combined algorithms in the No-AKI cohort.

**Table 4.**
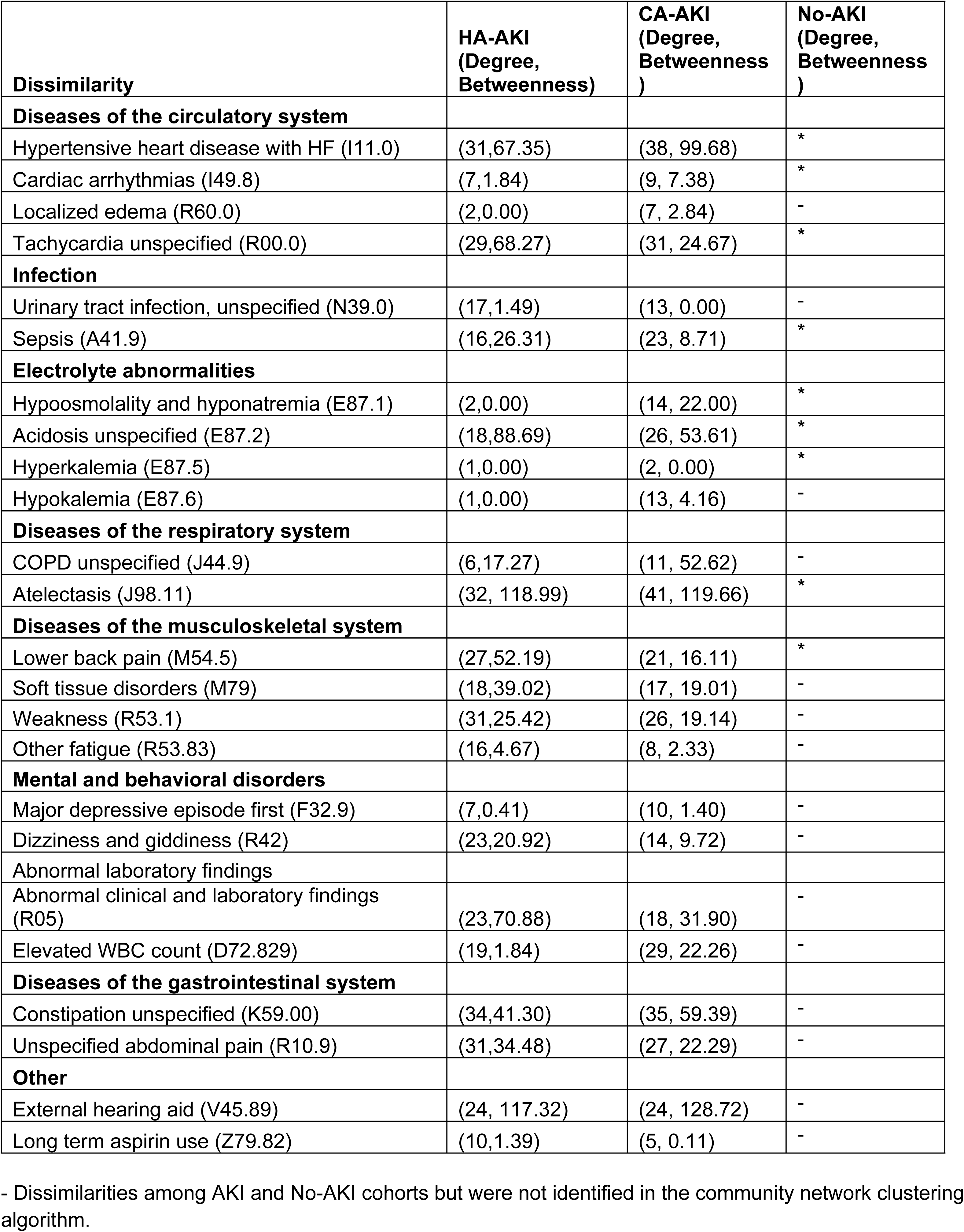
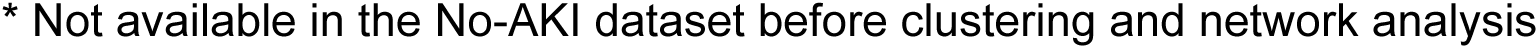
Dissimilarities of diagnosis among the cohorts.

| **Dissimilarity** | **HA-AKI**<br>**(Degree,**<br>**Betweenness)** | **CA-AKI**<br>**(Degree,**<br>**Betweenness**<br>**)** | **No-AKI**<br>**(Degree,**<br>**Betweenness**<br>**)** |
| --- | --- | --- | --- |
| **Diseases of the circulatory system** |  |  |  |
| Hypertensive heart disease with HF (I11.0) | (31,67.35) | (38, 99.68) | * |
| Cardiac arrhythmias (I49.8) | (7,1.84) | (9, 7.38) | * |
| Localized edema (R60.0) | (2,0.00) | (7, 2.84) | - |
| Tachycardia unspecified (R00.0) | (29,68.27) | (31, 24.67) | * |
| **Infection** |  |  |  |
| Urinary tract infection, unspecified (N39.0) | (17,1.49) | (13, 0.00) | - |
| Sepsis (A41.9) | (16,26.31) | (23, 8.71) | * |
| **Electrolyte abnormalities** |  |  |  |
| Hypoosmolality and hyponatremia (E87.1) | (2,0.00) | (14, 22.00) | * |
| Acidosis unspecified (E87.2) | (18,88.69) | (26, 53.61) | * |
| Hyperkalemia (E87.5) | (1,0.00) | (2, 0.00) | * |
| Hypokalemia (E87.6) | (1,0.00) | (13, 4.16) | - |
| **Diseases of the respiratory system** |  |  |  |
| COPD unspecified (J44.9) | (6,17.27) | (11, 52.62) | - |
| Atelectasis (J98.11) | (32, 118.99) | (41, 119.66) | * |
| **Diseases of the musculoskeletal system** |  |  |  |
| Lower back pain (M54.5) | (27,52.19) | (21, 16.11) | * |
| Soft tissue disorders (M79) | (18,39.02) | (17, 19.01) | - |
| Weakness (R53.1) | (31,25.42) | (26, 19.14) | - |
| Other fatigue (R53.83) | (16,4.67) | (8, 2.33) | - |
| **Mental and behavioral disorders** |  |  |  |
| Major depressive episode first (F32.9) | (7,0.41) | (10, 1.40) | - |
| Dizziness and giddiness (R42) | (23,20.92) | (14, 9.72) | - |
| Abnormal laboratory findings |  |  |  |
| Abnormal clinical and laboratory findings (R05) | (23,70.88) | (18, 31.90) | - |
| Elevated WBC count (D72.829) | (19,1.84) | (29, 22.26) | - |
| **Diseases of the gastrointestinal system** |  |  |  |
| Constipation unspecified (K59.00) | (34,41.30) | (35, 59.39) | - |
| Unspecified abdominal pain (R10.9) | (31,34.48) | (27, 22.29) | - |
| **Other** |  |  |  |
| External hearing aid (V45.89) | (24, 117.32) | (24, 128.72) | - |
| Long term aspirin use (Z79.82) | (10,1.39) | (5, 0.11) | - |

- Dissimilarities among AKI and No-AKI cohorts but were not identified in the community network clustering algorithm.
\* Not available in the No-AKI dataset before clustering and network analysis

Supplementary Table 4 highlights the diagnoses which were not selected by either algorithm across the three cohorts, indicating their uniqueness to a specific AKI cohort. For example, in the HA-AKI cohort, distinct diagnoses included hypertension (I95.9*, 6,1), dyspnea unspecified (14,25.06), pneumonia (4,5.16), diarrhea (16,35.79), nausea with vomiting (14,31.74) and dehydration (2,0). In contrast, in the No-AKI cohort, unique findings were family history of IHD (38, 94.38), breast cancer screening (23, 67.08), mammogram screening (11, 0.85), screening malignant neoplasm colon (19, 10.43) which were not identified in the AKI cohorts.

**Supplementary Table 4.** Unique phenotypes which were not matched by either algorithm.

Supplementary Table 5 shows the similarities in procedures across the three cohorts. Basic metabolic and electrolyte panels, along with levels of medical care services, are common across all three cohorts with similar centrality and betweenness values. Liver function tests varied among the cohorts. Specifically, HA-AKI had higher degree and betweenness values for aspartate aminotransferase enzyme(33, 19.84) compared to the other cohorts. In the CA-AKI cohort, alanine aminotransferase and alkaline phosphatase had higher betweenness values (27, 86.72) and (31, 45.31), respectively. The coagulation factor prothrombin time (PT) exhibited higher degree and betweenness value (34, 64.01) in the non-AKI cohort compared to the other two cohorts. Among cardiovascular serum laboratory values, natriuretic peptide and quantitative troponin had higher degree and betweenness values (34, 41.77; 30, 10.32) in the HA-AKI cohort relative to the others. In the microbiology category, all components except blood aerobic with isolation showed higher degree and betweenness in the HA-AKI cohort. For radiology, all imaging studies demonstrated higher degree and betweenness in the HA-AKI cohort.

**Supplementary Table 5.** Similarities of the procedures among the cohorts.

## 4. Discussion

### 4.1 Key findings

We investigated the occurrence of AKI in both community and hospital settings prior to a CKD diagnosis to identify key factors influencing future and sustained kidney damage. Early recognition and prevention are crucial for reducing CKD risk. Our network analysis corroborates known risk factors for AKI and CKD such as anemia, heart failure, and diabetes (Table 2) (38,39). Our algorithm revealed an 80% similarity in clinical phenotypes between the AKI (HA-AKI and CA-AKI) and non-AKI cohorts. A significant finding of our study is the identification of non-traditional comorbidities and additional risk factors for the progression of AKI to CKD, such as psychiatric disorders (e.g., major depressive disorder and anxiety) (Table 5). A 2021 population-matched cohort study of 30,998 patients with stress-related disorders (SRDs) found that patients with SRDs had an increased risk of AKI (HR 1.22, 95% CI 1.04-1.42) and CKD progression (HR 1.23, 95% CI 1.10-1.37) (Su et al., 2021). Our study supports these findings by identifying less commonly recognized risk factors for AKI and CKD progression through a clustering model (40,41). Although the No-AKI group had a higher comorbidity burden, our network clustering analysis revealed greater complexity in the comorbidity networks of both AKI cohorts compared to No-AKI cohort. This suggests that clinicians should consider the combined burden of risk factors rather than focusing on individual factors when managing AKI patients.

Historically the risk of AKI and its progression to CKD has been associated with factors such as increasing age, Black race, male gender, and the presence of multiple comorbidities (42–44).An observational study from 2021 suggests that the incidence of AKI increases after the age of 64 (45). However, our findings indicate that the AKI cohorts had a lower mean age compared to the non-AKI cohort and were predominantly White and female (Table 1). This suggests that AKI is not restricted to the elderly or any specific race or gender. The literature supports that older patients are at higher risk for AKI due to age-related declines in renal function and a higher number of comorbidities (46,47). Nonetheless, not all comorbidities contribute equally to the risk of AKI or CKD progression. Evaluating risk based solely on the number of comorbidities may not fully capture the associated risk. In our study, the non-AKI population exhibited a higher comorbidity burden compared to the AKI cohorts. These findings suggest that assessing the risk of AKI and its progression to CKD requires a comprehensive examination of various demographic and clinical factors to accurately identify specific risk factors and clinical phenotype hazards.

### 4.2 Network analysis comparison across three cohorts (HA-AKI, CA-AKI, no-AKI)

In our network analysis across the three cohorts, the AKI cohort exhibited higher quantitative values for edges and nodes compared to the non-AKI cohort, indicating a greater number of connections between nodes (Table 3). This analysis also revealed higher averages for degree and betweenness within the AKI cohort, particularly highlighting the overlap of comorbidities and demonstrating more closely clustered variables compared to the non-AKI cohort. Clinically, this suggests that the AKI cohort presents greater complexity, necessitating a more comprehensive examination of factors when assessing individuals at risk for CKD. Comorbidities shared across all cohorts were predominantly cardiac in nature, such as atrial fibrillation, heart failure, and atherosclerotic heart disease (Table 2). Conversely, sepsis, a well-known cause of AKI in critically ill patients and a major contributor to ICU morbidity and mortality, was highlighted. A 2011 study found that 40% of critically ill patients develop sepsis following AKI, indicating that AKI may increase the risk of sepsis (48). The challenge of determining which syndrome occurs first is notable, as both conditions are interrelated. Our findings align with previous research, particularly in comparing HA-AKI to CA-AKI cohorts. We observed that sepsis had higher betweenness in the HA-AKI cohort (26.31 vs. 8.71), underscoring its significant association with AKI risk (Table 4). Additionally, diagnoses leading to intravascular volume depletion, such as diarrhea, nausea with vomiting, and dehydration, were uniquely associated with the HA-AKI cohort. These results support existing literature and further emphasize the importance of these phenotypes in identifying HA-AKI.

### 4.3 Similarities and Dissimilarities of procedures among cohorts

Our analysis revealed that patients in the AKI cohorts had higher degrees and betweenness for procedures related to basic metabolic and electrolyte panels. This is consistent with expectations, as patients with kidney disease typically require closer monitoring of serum electrolyte levels (47). In the HA-AKI cohort, aspartate aminotransferase enzyme measurements showed higher degrees and betweenness compared to the other cohorts. While the development of CKD post-AKI and its associated risk factors are well-documented in patients without cirrhosis (49), our study did not include patients with cirrhosis. The increased degree and betweenness of aspartate aminotransferase in the HA-AKI cohort may suggest potential hepatic involvement, despite the absence of cirrhosis. Conversely, in the non-AKI cohort, cardiovascular serum laboratory values such as natriuretic peptide and quantitative troponin levels exhibited higher degrees and betweenness (34, 41.77; 30, 10.32) compared to the AKI cohorts (13, 4.71; 11, 0.91 and 13, 3.42; 6, 0.37, respectively). This finding is contrary to expectations, given the established link between AKI and cardiovascular disease (50). This discrepancy highlights an area that warrants further investigation to reconcile these unexpected results.

### 4.4. Custom combination of clustering and network-based methods

Unlike methods that focus solely on grouping individuals, ClustOfVar analyzes the data structure by clustering variables based on their intrinsic relationships. It accommodates both quantitative and qualitative variables, creating synthetic variables to reduce complexity while preserving essential information. ClustOfVar enhances intra-cluster cohesion through a homogeneity criterion, a feature not commonly emphasized in other methods. This approach provides a more nuanced understanding of data complexity, uncovering hidden patterns and dependencies that might be missed in patient-level analyses. The hierarchical clustering routine and its dendrogram facilitate visual interpretation and the determination of the optimal number of clusters, making it a robust tool for analyzing diverse and complex datasets.

### 4.5 Strengths and implications of the proposed method

Our innovative comprehensive patient profiling tool accurately projects a patient’s progression from AKI to CKD using diagnosis and procedures data. This custom profiling algorithm integrates two clustering methods (ClustOfVar and network-based community clustering), thereby enhancing the validity of traditional clustering approaches. Our network analysis not only quantifies the number of comorbidities and procedures associated with AKI to CKD progression but also identifies specific types of comorbidities and procedures relevant to the patient’s journey. This tool confirms established risk factors for AKI to CKD progression, such as hypertension and diabetes, and further highlights non-traditional risk factors, including alcohol use disorder and characteristics specific to the non-AKI group. This tool can be applied to understand the progression of other chronic diseases (e.g., heart failure, COPD, diabetes). For instance, it demonstrates how different comorbidities and procedures can be analyzed using our profiling method to elucidate factors influencing the transition from AKI to CKD. It identifies both similar and distinct risk factor clusters among patient groups. Unlike previous studies that have focused on pre-selected risk factors (e.g., diabetes and hypertension), our tool provides a comprehensive list of otherwise unidentified risk factors affecting AKI to CKD progression. Additionally, it can be used to assess the long-term impact of chronic diseases, particularly by linking the clusters revealed in both clustering and network analyses to demographic factors such as age, gender, race, and ethnicity.

### 4.6. Limitations

Our study has several limitations that should be considered. First, the retrospective design introduces the risk of missing data, which could lead to confounding bias. For instance, AKI and CKD patients were identified using ICD-9 and ICD-10 codes due to the absence of serum creatinine values. Second, our definition of CA-AKI was based on inclusion and exclusion criteria, as we could not measure prior exposure to CA-AKI. Third, the results may be affected by residual biases and unmeasured confounders, as some commonly associated conditions (e.g., diseases or procedures with <5% prevalence) were excluded. Fourth, our algorithm does not capture temporal relationships between comorbidities and procedures related to AKI progression to CKD, largely due to gaps in continuous healthcare insurance coverage.

## 5. Conclusion

Specific therapeutic strategies to prevent the progression from AKI to CKD are currently limited. Our custom comprehensive patient profiling algorithm offers a method for grouping and identifying phenotypes of AKI patients based on their comorbidities and medical procedures. This approach holds promise for advancing research in large cohorts or electronic health records, enabling a deeper understanding of various AKI phenotypes and addressing the clinical gap leading to CKD. Future research should include prospective, multi-center studies to evaluate the impact of different stages and durations of AKI on the risk of long-term CKD complications. Additionally, exploring temporal patterns of comorbidities and procedures in relation to other chronic diseases using Medicaid and private healthcare claims data could be beneficial. This tool also has the potential to uncover non-traditional risk factors for chronic diseases by identifying less prevalent or previously unknown comorbidities and procedures.

## Data Availability

The dataset could be requested to TriNetX upon an agreement with the requester.

